# Neutralisation of the SARS-CoV-2 Delta sub-lineage AY.4.2 and B.1.617.2 + E484K by BNT162b2 mRNA vaccine-elicited sera

**DOI:** 10.1101/2021.11.08.21266075

**Authors:** Ria Lassaunière, Charlotta Polacek, Jannik Fonager, Marc Bennedbæk, Lasse Boding, Morten Rasmussen, Anders Fomsgaard

## Abstract

Several factors may account for the recent increased spread of the SARS-CoV-2 Delta sublineage AY.4.2 in the United Kingdom, Romania, Poland, and Denmark. Here, we evaluate the sensitivity of AY.4.2 to neutralisation by sera from Pfizer/BioNTech (BNT162b2) vaccine recipients. AY.4.2 neutralisation was comparable to other circulating Delta lineages or sublineages. In contrast, the more rare B.1.617.2+E484K variant showed a significant >4-fold reduction in neutralisation that warrants surveillance of strains with the acquired E484K mutation.

## Background

In recent weeks, the SARS-CoV-2 Delta sublineage AY.4.2 has accounted for an increased proportion of Delta cases in the United Kingdom (UK), rising from 3.8% to 11.3% in the weeks beginning with the 19^th^ of September 2021 and the 18^th^ of October 2021, respectively [1, 2]. Multiple other countries reported its presence with over 26 000 AY.4.2 sequences from 42 countries uploaded on GISAID to date [3], and frequencies above 1% observed in Romania and Poland [4]. On the 20^th^ of October 2021, the UK Health Security Agency designated AY.4.2 a Variant Under Investigation (VUI-21OCT-01) as it appears to have a higher growth rate (19%) in the population relative to other Delta lineages and sublineages [1]. AY.4.2 bears the same spike mutations as AY.4 with the addition of Y145H and A222V. Neutralisation data for AY.4.2 is currently lacking and warrants rapid evaluation to establish if neutralisation resistance may account for the increased spread of this sublineage in populations, where a majority has been vaccinated. We further evaluate neutralisation of a Delta strain bearing the known neutralisation resistant mutation, E484K [5, 6]. The latter amino substitution has sporadically occurred in Delta strains, but recently established small clusters in the UK [2] and contributed to breakthrough infections in Italy [7]. Here we investigate the sensitivity of AY.4.2 and the Delta lineage B.1.617+E484K to neutralisation by SARS-CoV-2 vaccine-induced anti-sera.

### Cohort

We assessed the SARS-CoV-2 Delta sublineage AY.4.2 virus neutralisation using serum samples from BNT162b2 (Pfizer/BioNTech) mRNA vaccinated individuals (N = 30) between the ages of 20 and 91 years (median: 42 years; interquartile range: 35 to 63 years); 23.3% were male [8]. The time between the second vaccination and sampling ranged from 30 to 100 days (median: 61 days; interquartile range: 51 to 64 days). All donors developed anti-SARS-CoV-2 spike antibodies after vaccination as determined by the Wantai Total Ab ELISA assay [9, 10]. The samples were excess material from diagnostic testing conducted at Statens Serum Institut, Denmark. Samples were taken for diagnostic purposes unrelated to COVID-19; however, are linked to the Danish Vaccination Registry that enables surveillance of immune escape by vaccine-induced antibody responses.

### Neutralisation assay

Virus neutralisation was tested against SARS-CoV-2 clinical isolates cultured on Vero E6 cells (Table 1). All virus stocks were sequenced to confirm the presence of lineage-specific mutations and the absence of cell culture-derived mutations. Inhibition of live virus infection is measured in an anti-SARS-CoV-2 nucleocapsid protein ELISA. The ELISA signal was not affected by amino acid substitutions present in the nucleocapsid protein of one or more of the isolates. The quantitative optical density values from the two-fold serial dilution of serum (range: 1:10 to 1:1280) enables calculation of exact 50% neutralisation titers using four-parameter logistic regression. The assay has a comparable performance relative to other neutralisation assays used in different European laboratories (laboratory 4 in ref [11]). Titers were not normally distributed (Shapiro-Wilk test) and were compared using the nonparametric Friedman test for paired measurements followed by Dunn’s multiple comparison test. Adjusted P-values are reported.

**Table 1.** SARS-CoV-2 viral isolates evaluated for neutralisation using BNT162b2 vaccine sera.

| Variant | Pango lineage | Strain |
| --- | --- | --- |
| Early pandemic (D614G) | B.1 | SARS-CoV-2/Hu/Denmark/SSI-H1 |
| Alpha | B.1.1.7 | SARS-CoV-2/Hu/Denmark/SSI-H14 |
| Beta | B.1.351 | hCoV-19/Netherlands/NoordHolland_10159/2021 |
| Gamma | P.1 | SARS-CoV-2/Hu/Denmark/SSI-H26 |
| Delta | B.1.617.2 | SARS-CoV-2/Hu/Denmark/SSI-H11 |
| Delta | AY.4 | SARS-CoV-2/Hu/Denmark/SSI-H38 |
| Delta | AY.4.2 | SARS-CoV-2/Hu/Denmark/SSI-H37 |
| Delta | B.1.617.2+E484K | SARS-CoV-2/Hu/Denmark/SSI-H41 |

### AY.4.2 virus neutralisation relative to other Delta variants

We first assessed neutralisation of the AY.4.2 virus by vaccine sera relative to other Delta strains bearing lineage and sublineage-defining spike protein amino acid changes as well as an early pandemic strain with the single spike D614G amino acid substitution (Figure 1). All tests were performed concurrently to ensure minimal inter-assay variation. Relative to the early pandemic strain (D614G), the AY.4.2 virus had a 2.3-fold reduction in median neutralisation titers (median titer: 199 vs. 87; P < 0.0001) (Figure 2). However, the titers for AY.4.2 did not differ significantly from those measured for the parental Delta lineage B.1.617.2 (median titer: 87 vs. 118; P > 0.050) or Delta sublineage AY.4 (median titer: 87 vs. 118; P > 0.050). We further evaluated a B.1.617.2 lineage isolate that contains the E484K amino acid substitution in the receptor-binding domain. In contrast to AY.4.2, the B.1.617.2 strain with E484K had a significant reduction in virus neutralisation titers relative to D614G (4.0-fold) and all other Delta strains tested – B.1.617.2 (2.3-fold), AY.4 (2.3-fold), and AY.4.2 (1.7-fold) (P < 0.050 for all comparisons).

**Figure 1.**
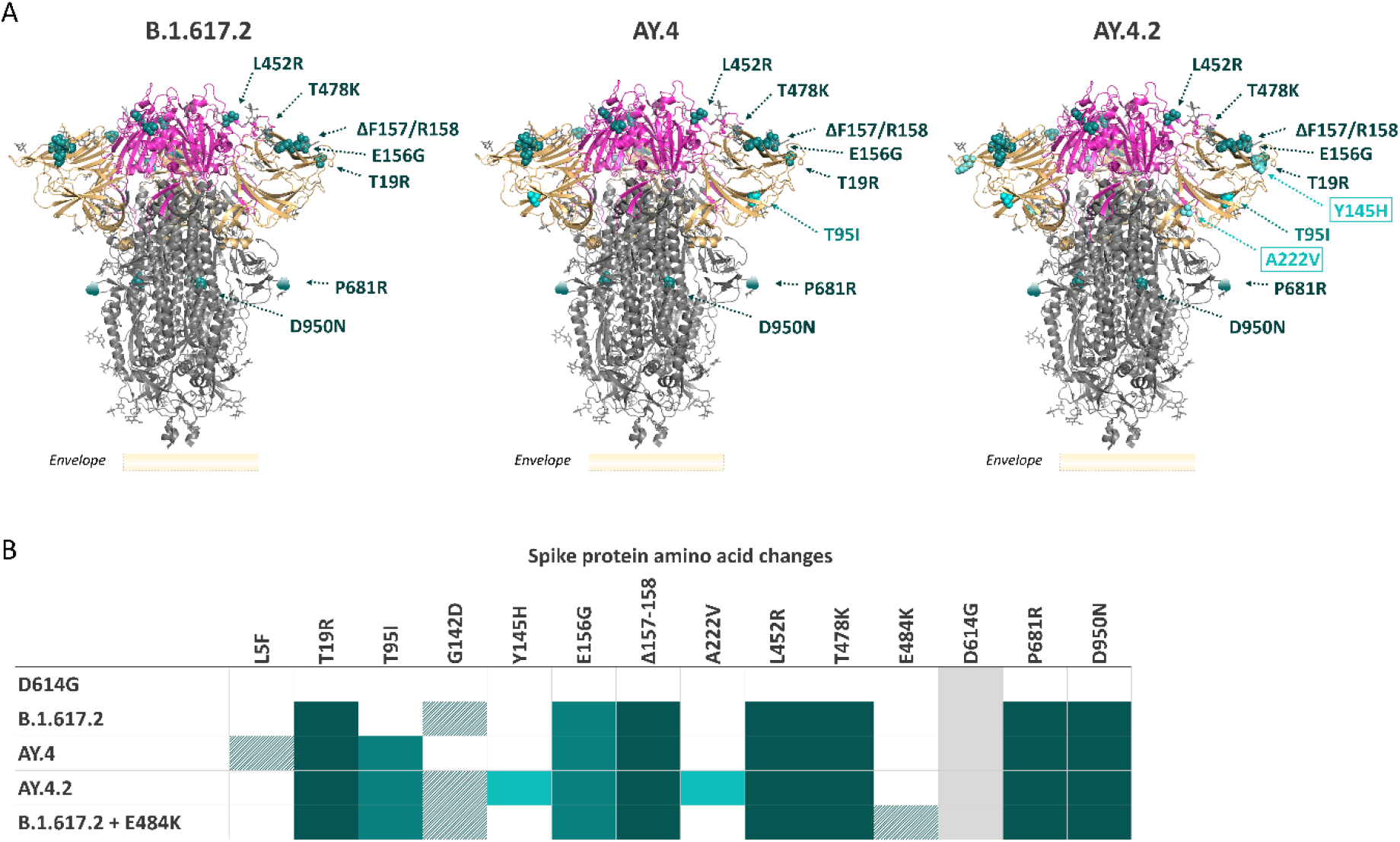
Relative positions of spike mutations of SARS-CoV-2 Delta lineage B.1.617.2, AY.4 and AY.4.2. A) Pre-fusion SARS-CoV-2 spike crystal structure with receptor binding domain in magenta, N-terminal domain in beige, S2 domain in grey, and amino acid substitutions and deletions relative to the Wuhan-Hu-1 reference sequence indicated by cyan spheres. Dark turquoise indicates B.1.617.2 lineage mutations, light turquoise indicates AY.4 sub-lineage mutations, and light cyan indicates AY.4.2 sub-lineage mutations. B) Spike mutations of SARS-CoV-2 clinical isolates used to evaluate virus neutralisation of an early pandemic strain D614G, B.1.617.2, AY.4, AY.4.2, and B.1.617.2 + E484K.

**Figure 2.**
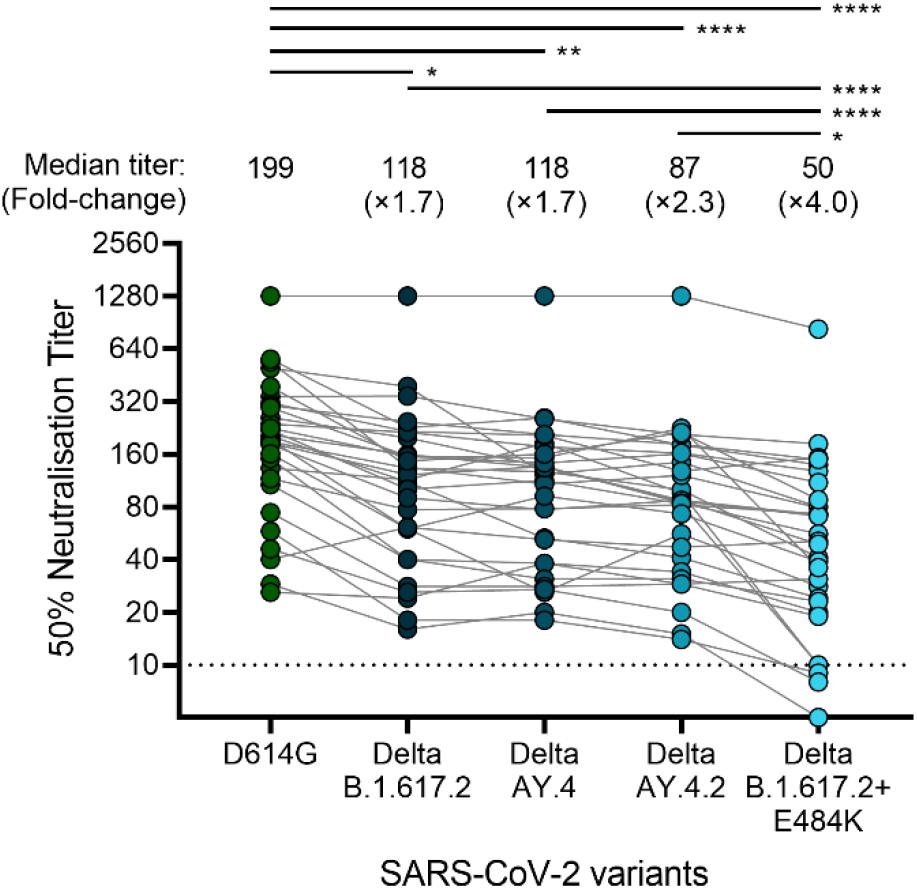
SARS-CoV-2 neutralising antibody titers against Delta sublineage AY.4.2 after the second dose of BNT162b2 mRNA vaccination. Virus neutralisation for each serum samples was tested against clinical isolates representing an early pandemic strain (D614G), the parental Delta lineage B.1.617.2, Delta lineage AY.4, Delta sublineage AY.4.2, and a Delta lineage B.1.617.2 bearing the neutralisation resistant spike mutation E484K. Light grey lines connect data from each sample for the different virus isolates. N = 30 serum samples, each taken from a different individual. Dotted line, limit of detection (<10). *P<0.05, **P<0.01, ****P<0.0001

### AY.4.2 virus neutralisation relative to variants of concern (VOC)

Virus neutralisation titers for other VOC were available for 24 of the 30 serum samples (Figure 3). Relative to the early pandemic strain (D614G), the reduction in the AY.4.2 sublineage-associated virus neutralisation (2.3-fold) was not as pronounced as observed for the Beta variant (4.9-fold). In contrast, the Delta lineage B.1.617.2 with the E484K neutralisation resistant mutation (4.4-fold) approached the reduction in neutralisation titers observed for the Beta variant.

**Figure 3.**
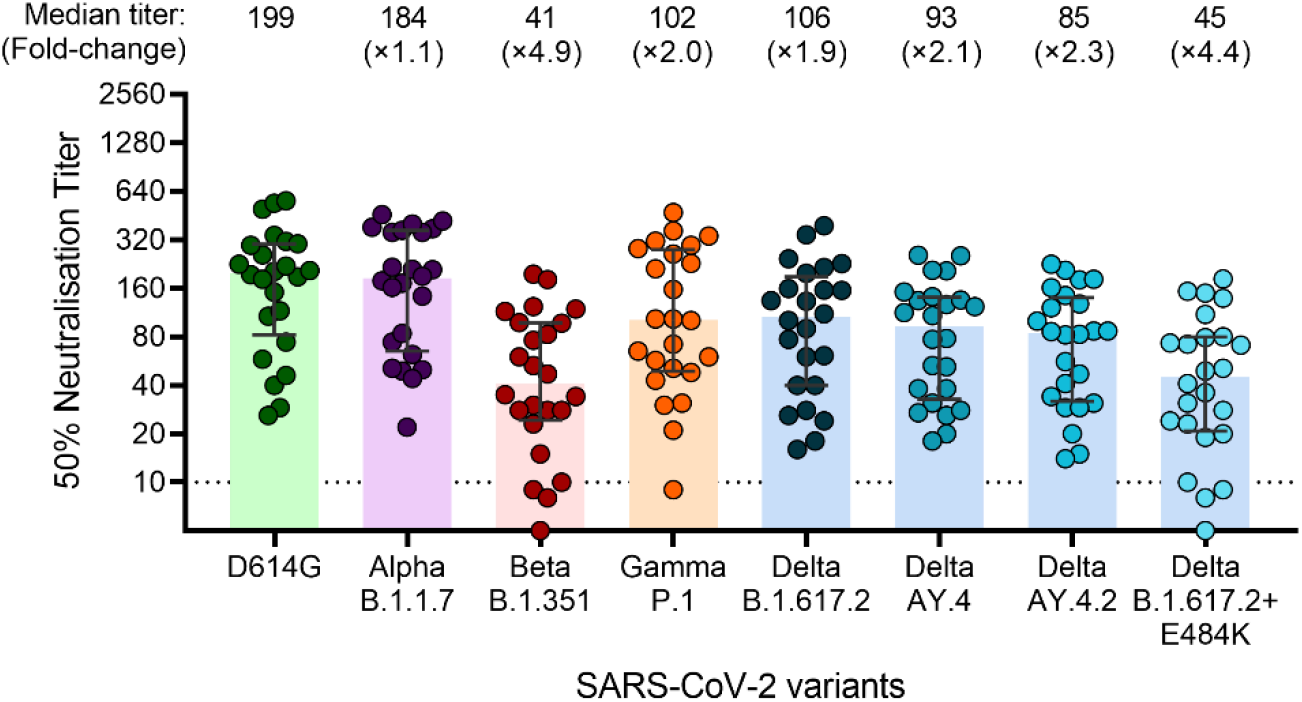
SARS-CoV-2 neutralising antibody titers against variants of concern Alpha, Beta, Gamma and Delta lineage/sub-lineages B.1.617.2, AY.4, AY.4.2, and B.1.617.2+E484K two months after the second dose of BNT162b2 mRNA vaccination. Neutralisation titers for variants of concern Alpha, Beta and Gamma in addition to the Delta variant strains were available for 24 of the 30 vaccinated sera. Bars represent the median titer and error bars the interquartile range; N = 24.

### Prevalence of AY.4.2. and B.1.617.2 with E484K in Denmark

In Denmark, the first AY.4.2 case was observed on the 4^th^ August 2021. In September 2021, AY4.2 appeared in more than 2% of sequenced samples but declined to below 1%. However, during the last week of October 2021, AY.4.2 increased again from 0.5% to 2%. Beyond Beta and Gamma, the E484K mutation has emerged in several different lineages, including Delta [7, 12]. To date, 37 cases of Delta strains with the E484K mutation has been identified in Denmark between the 29^th^ of August 2021 and the 5^th^ of November 2021.

## Discussion and conclusions

In addition to the spike mutations present in the prevalent AY.4 sublineage, AY.4.2 bears the Y145H and A222V amino acid substitutions. Both occur in the N-terminal domain of the spike protein. A222V has been observed in earlier emerging SARS-CoV-2 variants, but is not considered to contribute to increased transmissibility or immune escape in those variants [13]. A functional consequence for Y145H remains to be established. Using a live virus neutralisation assay, we demonstrate that the Delta sublineage AY.4.2 virus has a modest reduction of 2.3-fold relative to an early pandemic strain, which is highly homologous to the current vaccine strains. AY.4.2 therefore remains sensitive to vaccine-induced virus neutralisation. Moreover, neutralisation titers for AY.4.2 were not significantly different from the parental B.1.617.2 or AY.4 lineage. It is thus unlikely that neutralisation resistance is a determinant of the increased spread observed for AY.4.2 relative to other Delta lineages in European countries. These findings are in agreement with a similar vaccine effectiveness observed for AY.4.2 compared to non-AY.4.2 Delta cases, both symptomatic and asymptomatic, for the Astra Zeneca, Pfizer/BioNTech and Moderna vaccines in the UK [2]. On the contrary, we show a significant neutralisation resistance for a Delta variant that acquired the E484K spike mutation. This amino acid substitution in the receptor-binding domain, occur in other VOC that include Beta and Gamma with reduced sensitivity to monoclonal antibodies, convalescent antisera from early pandemic wave infections, and vaccine elicited anti-sera [6, 14, 15]. While Delta variants bearing the latter mutation occur mostly sporadically [7], it has now occurred in Delta sublineages with sustained clusters recently reported in the UK [2]. The presented data provide further support for continuous monitoring of E484K within emerging Delta sublineages such as the Delta strain examined here.

## Data Availability

All data produced in the present work are contained in the manuscript

## Conflict of interest

None declared

## Funding statement

No particular funding was obtained for this work, which was a part of the Danish national health response to the COVID-19 pandemic.

## Ethical statement

The study was conducted according to the Declaration of Helsinki. Biological samples were stored as part of the Statens Serum Institut’s diagnostic testing and surveillance. Ethical approval was not required for this surveillance study.

## Notes

### Competing Interest Statement

The authors have declared no competing interest.

### Author Declarations

Exemption for review by the ethical committee system and informed consent was given by the Committee on Biomedical Research Ethics - Capital Region in accordance with Danish law on surveillance projects.

### Summary of Updates

Increased sample number

